# Interactive network-based clustering and investigation of multimorbidity association matrices with associationSubgraphs

**DOI:** 10.1101/2022.02.17.22271014

**Authors:** Nick Strayer, Siwei Zhang, Lydia Yao, Tess Vessels, Cosmin A Bejan, Ryan S Hsi, Jana K Shirey-Rice, Justin M Balko, Douglas B Johnson, Elizabeth J Philips, Alex Bick, Todd L Edwards, Digna R Velez Edwards, Jill M Pulley, Quinn S Wells, Michael R Savona, Nancy J Cox, Dan M Roden, Douglas M Ruderfer, Yaomin Xu

**Affiliations:** Department of Biostatistics, Vanderbilt University, Nashville, TN, USA; Vanderbilt Genetics Institute, Vanderbilt University Medical Center, Nashville, TN, USA; Department of Biomedical informatics, Vanderbilt University Medical Center, Nashville, TN, USA; Department of Urology, Vanderbilt University Medical Center, Nashville, TN, USA; Vanderbilt Institute for Clinical and Translational Research, Vanderbilt University Medical Center, Nashville, TN, USA; Division of Hematology and Oncology, Department of Medicine, Vanderbilt University Medical Center, Nashville, TN, USA; Center for Drug Safety and Immunology, Department of Medicine, Vanderbilt University Medical Center, Nashville, TN, USA; Institute for Immunology and Infectious Diseases, Murdoch University, Murdoch, Western Australia, Australia; Division of Genetic Medicine, Department of Medicine, Vanderbilt University Medical Center, Nashville, TN, USA; Division of Epidemiology, Department of Medicine, Vanderbilt University Medical Center, Nashville, TN, USA; Department of Obstetrics and Gynecology, Vanderbilt University Medical Center, Nashville, TN, USA; Department of Allergy, Pulmonary and Critical Care Medicine, Vanderbilt University School of Medicine, Nashville, TN, USA; Department of Cardiovascular Medicine, Vanderbilt University Medical Center, Nashville, TN, USA; Department of Internal Medicine, Vanderbilt University Medical Center, Nashville, TN, USA; Department of Pharmacology, Vanderbilt University Medical Center, Nashville, TN, USA; Department of Psychiatry and Behavioral Sciences, Vanderbilt University Medical Center, Nashville, TN, USA

## Abstract

**Summary:** Making sense of association networks is vitally important to many areas of high-dimensional analysis. Unfortunately, as the data-space dimensions grow, the number of association pairs increases in *O*(*n*^*2*^); this means traditional visualizations such as heatmaps quickly become too complicated to parse effectively. Here we present associationSubgraphs: a new interactive visualization method to quickly and intuitively explore high-dimensional association datasets using network science-derived statistics and visualization. As a use case example, we apply associationSubgraphs to a phenome-wide multimorbidity association matrix generated from an electronic health record (EHR) and provided an online, interactive demonstration for exploring multimorbidity subgraphs.

**Availability:** An R package implementing both algorithm and visualization components of associationSubgraphs is available at https://github.com/tbilab/associationsubgraphs. Online documentation is available at https://prod.tbilab.org/associationsubgraphs_info/. A demo using a multimorbidity association matrix is available at https://prod.tbilab.org/associationsubgraphs-example/.

## 1 INTRODUCTION

Analysis of association or correlations between variables is a very important step in exploratory data analysis of high-dimensional datasets. In these scenarios, a dataset with a large number of columns or measured variables but without known or validated patterns of association among them is inspected using statistical and visualization methods to gain insight into how the variables may interact with each other. There are many different ways of establishing the strength of these interactions, from as simple as the mutual occurrence of binary variables (Cha, 2007), to complex penalized regression models (Hallac *et al*., 2015; Tibshirani *et al*., 2005). Examples of areas where association analysis is used include gene regulatory networks (Gustafsson *et al*., 2005), analysis of single-cell sequencing data to determine cell differentiation (Chan *et al*., 2017), networks of comorbidity between diseases (Chen and Xu, 2014), topic modeling in natural language processing (Wang and Zhu, 2014).

Traditional analysis of these association patterns uses heatmaps. In these visualizations, both rows and columns represent all present variables, and the color of the cells represents the strength of the association between the two variables. As the number of variables grows larger the effectiveness of heatmap rapidly decreases. One important issue is the ordering of the rows and columns, as the precise placement of a variable in relation to other completely change inference made by the analyst(Bojko, 2009) and thus must be done carefully. Typically, this ordering is done by a clustering algorithm(Metsalu and Vilo, 2015; Pryke *et al*., 2007), which injects model assumptions into the visualization that are not immediately clear to the analyst or later audience. Another issue with large heatmaps is the difficulty of discerning the identity of cells that fall far from the labeled axes(Bojko, 2009).

One way of alleviating the issues with heatmaps is to reduce the association space using edge or association filtering and then visualize the results using network visualization tools. These methods typically involve parametric tests that do not account for the network structure (Benjamini and Yekutieli, 2001) or contain many assumptions (Hallac *et al*., 2015). Non-parametric methods typically based on permutations also exist but, due to the *O*(*n*^*2*^) complexity inherent in association datasets, are computationally infeasible for large datasets (Harris and Drton, 2013). We point the reader to the book by Kolaczyk and Csárdi, 2020 for more information on these methods.

A classic model often studied in network science is the “random graph” (Solomonoff & Rapoport, 1951) (often called the Erdos-Renyi graph). In random graphs, nodes are connected by a given number of edges randomly, i.e., without preference for specific nodes. An emergent property of these “random” graphs are “components” or “isolated subgraphs” (Mark Newman, 2018). Isolated subgraphs are groups of nodes connected internally but not to any other nodes in the network. Percolation theory (Mark Newman, 2018) is a subfield of network science dedicated to understanding how the removal of edges within a network leads to the formation of these isolated subgraphs.

We previously developed PheWAS-ME: an interactive dashboard to visualize individual-level genotype and phenotype data side-by-side with PheWAS analysis results, allowing researchers to explore multimorbidity patterns and their associations with a genetic variant of interest(Strayer *et al*., 2021). In this work, by framing association analysis as a network problem, we can utilize the results of percolation theory to design an intuitive set of visualizations for exploring the subgraph structure of multimorbidity association networks based around the concept of adding and removing edges in the order of the strength of association. We name this algorithm associationSubgraphs and implement it as a interactive visualization method to quickly explore high-dimensional association datasets. As a use case example, we applied associationSubgraphs to a phenome-wide multimorbidity association matrix generated from an electronic health record (EHR) and provide an online, interactive demonstration for exploring multimorbidity subgraphs.

## 2 METHODS

### 2.1 Algorithm

The algorithm for computing associationSubgraphs at all given cutoffs is closely related to single-linkage clustering (Gower and Ross, 1969) but differs philosophically by viewing nodes that are yet to be merged with other nodes as unclustered rather than residing within their own cluster of size one.

To calculate the set of subgraphs at every threshold the algorithm starts by sorting edges/associations in descending order of strength. Then, the nodes connected by the highest association strength are set as a “cluster”. Next, the second-highest association strength is added. If either adjacent node is shared with the first cluster, the non-shared node is added to the existing cluster. Otherwise, a new separate cluster containing the two adjacent nodes is created. This procedure is repeated for all association pairs. If both nodes for a pair already reside in separate clusters, then those clusters are merged into a new “super” cluster. After every edge is added, the current cluster state is exported. For further details, see Algorithm 1.

### 2.2 Visualization

The subgraph-clustering algorithm results are visualized through an interactive visualization built using the javascript library D3 (Bostock *et al*., 2011) that allows panning through and visualizing all cluster states that occur during the running of the algorithm.

At every step, all currently clustered nodes are displayed as a grid of force-directed subgraphs (de Leeuw, 1988) (Figure 1A). Accompanying each subgraph is a set of three measures as encoded in a chart (Figure 1B). These are the number of nodes in the subgraph, the density (number of edges at current threshold linking nodes relative to maximum possible), and the average strength of all those edges.

**Figure 1:**
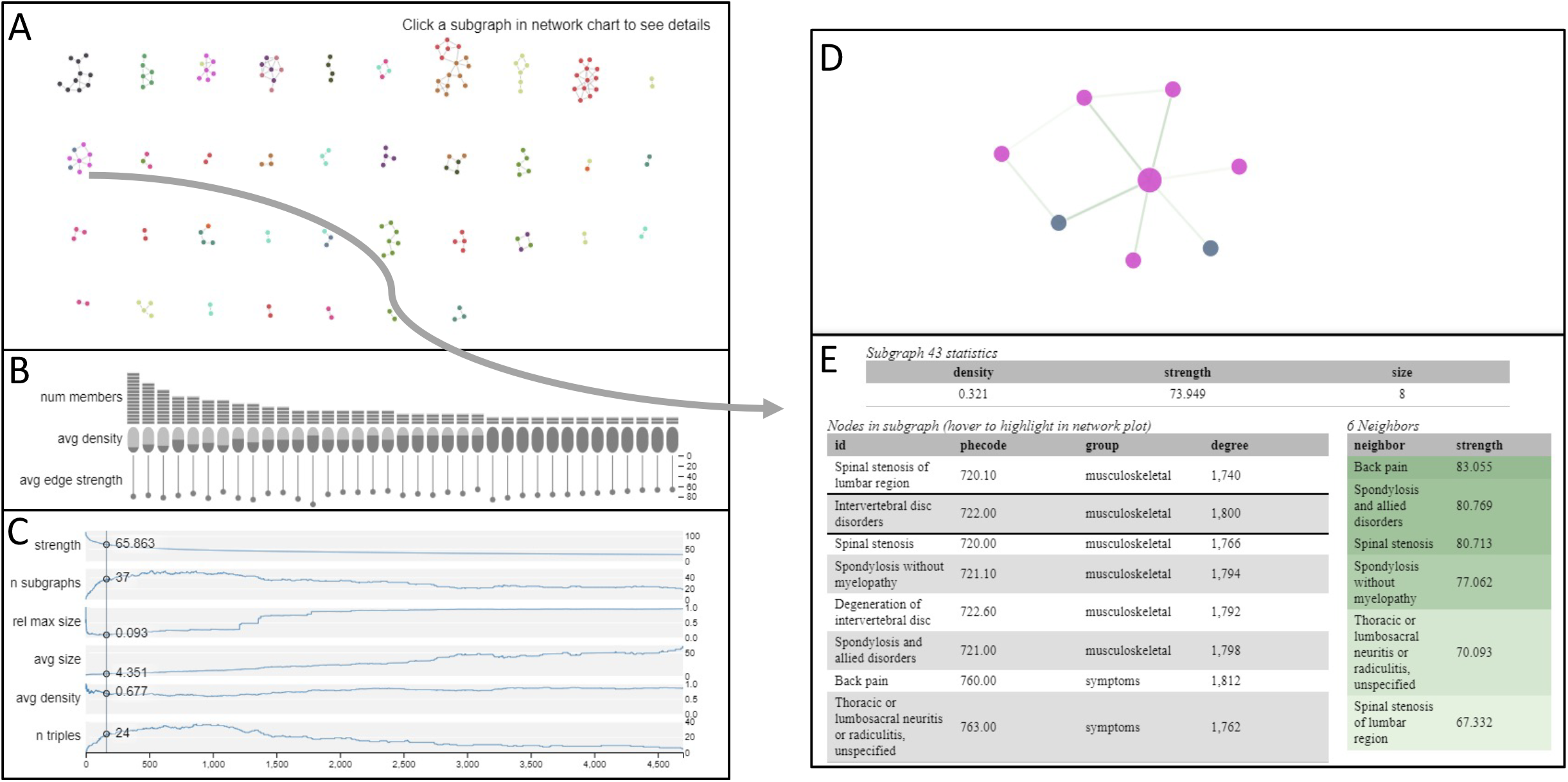
Interactive visualization of subgraph clustering results with current threshold set at the optimal threshold according to the smallest-largest rule

By adding edges in one-at-a-time, we are emulating the formation of a random graph. As the edges are added, isolated subgraphs form within the network. By separating the subgraphs, the visualization acknowledges that, at the current association strength threshold, the nodes are conceptually isolated and should be represented as such, unlike traditional network or heat map visualizations.

To aid in the selection of association threshold, a series of line-plots below the network visualization (Figure 1C) provide summary statistics about the cluster state at every possible cutoff. These include the number of subgraphs, the number of subgraphs with at least three members (triples), the average density of those subgraphs, the average size of the subgraphs, the size of the largest subgraph relative to all other subgraphs combined, and the current association threshold. By hovering over a state in these line plots the drawn network updates to represent the desired threshold. This updating is done in real-time allowing the user to see exactly what edges were added and how those edges changed the subgraph state.

Every resulting subgraph can be selected and zoomed into by clicking, which reveals all members within the cluster (Figure 1D), the edge strengths between them, and any further supplementary node information provided by the user (Figure 1E).

### 2.3 Choosing “optimal” threshold

AssociationSubgraphs is meant to provide an exploratory view of an entire association network; this means the concept of the ideal threshold is not particularly important. However, we can draw inspiration from random-graph and percolation theory to provide an estimate of an “optimal” threshold value used as the initial point in the visualization.

When nodes are truly randomly connected together, what is known as a “giant component” form very quickly. A giant component is one in which a very large portion, typically n^2/3^ (Mark Newman, 2018), of the nodes in the network are in the same isolated subgraph. There are three “phase-transitions” regarding the size of the largest isolated subgraph in the network relative to the number of edges (*e*) added.

If *e*<*n*, we would expect many small subgraphs with the largest size on the order of log(*n*). If *e*=*n*, then we would expect to still have a large number of subgraphs, with an expected largest subgraph of size n^2/3^. Last, if *e*>*n* we would expect all nodes to be connected in one giant subgraph/component.

When the edges are not purely random, we expect deviation from these patterns, and in practice we see these deviations; with a giant component typically forming well after the number of included edges surpasses the number of nodes.

To take advantage of this known behavior, we propose the “largest-smallest” rule for finding the optimal threshold. This rule states that the optimal threshold value for an association network will be just before the giant component starts to form. This point is estimated by tracking the aforementioned size of the largest subgraph relative to all other subgraphs metric. When this metric starts to rise, it indicates edges are now being added mostly at random; thus, the optimal threshold can be seen as the minimum of this function.

### 2.4 Example Usage

Multimorbidity, defined as the co-existence of two or more concurrent health conditions in one patient, can be represented as networks with diseases as nodes and their connections as links, typically weighted according to the strength of pair-wise disease associations. Figure 1 shows the results of running the associationSubgraphs algorithm and visualization on such a multimorbidity network of 1,815 phenotypes as “Phecodes” (Denny *et al*., 2010) constructed using Vanderbilt EHR data. The network strength measurements are based on the test statistics of a regression analysis assessing the association between each Phecode pairs. AssociationSubgraphs provides intuitive and meaningful insights into the subgraph structure of the example multimorbidity network. Using the described smallest-largest point to determine an association strength cutoff returns a network with 36 isolated subgraphs. Figure 1E shows the investigation of one of the present subgraphs including the codes 720.00, 720.10, 721.00, 721.10, 722.00, 722.60, 760.00, 763.00, which are all back pain related Phecodes (e.g., Spinal stenosis of the lumbar region: 720.10, and Back pain: 760.00.) More examples and this particular visualization are available on the associationSubgraphs R package website (see availability).

## 3 CONCLUSIONS

In this paper we have provided a brief introduction to the algorithm and visualization associationSubgraphs for exploring association networks. By enabling the exploration of high-dimensional association networks through interactive network visualization guided by basic network-science theory, associationSubgraphs allows researchers to understand their association network data with greater precision and intuition. We applied associationSubgraphs to a large-scale disease multimorbidity association matrix generated from EHR and provided an online, interactive demo for exploring multimorbidity subgraphs.

## Data Availability

An R package implementing both algorithm and visualization components of associationSubgraphs is available at https://github.com/tbilab/associationsubgraphs. Online documentation is available at https://prod.tbilab.org/associationsubgraphs_info/. A demo using a multimorbidity association matrix is available at https://prod.tbilab.org/associationsubgraphs-example/.

https://prod.tbilab.org/associationsubgraphs_info/

https://prod.tbilab.org/associationsubgraphs-example/

## Author Declarations

All relevant ethical guidelines have been followed and any necessary IRB and/or ethics committee approvals have been obtained.

Yes

All necessary patient/participant consent has been obtained and the appropriate institutional forms have been archived.

Yes

Any clinical trials involved have been registered with an ICMJE-approved registry such as ClinicalTrials.gov and the trial ID is included in the manuscript.

Not Applicable

I have followed all appropriate research reporting guidelines and uploaded the relevant Equator, ICMJE or other checklist(s) as supplementary files, if applicable.

Yes

### Algorithm 1: Find neighborhood subgraphs

*By simply requiring the association strengths to be sorted, the only assumption required of the strength measure is monotonicity*.

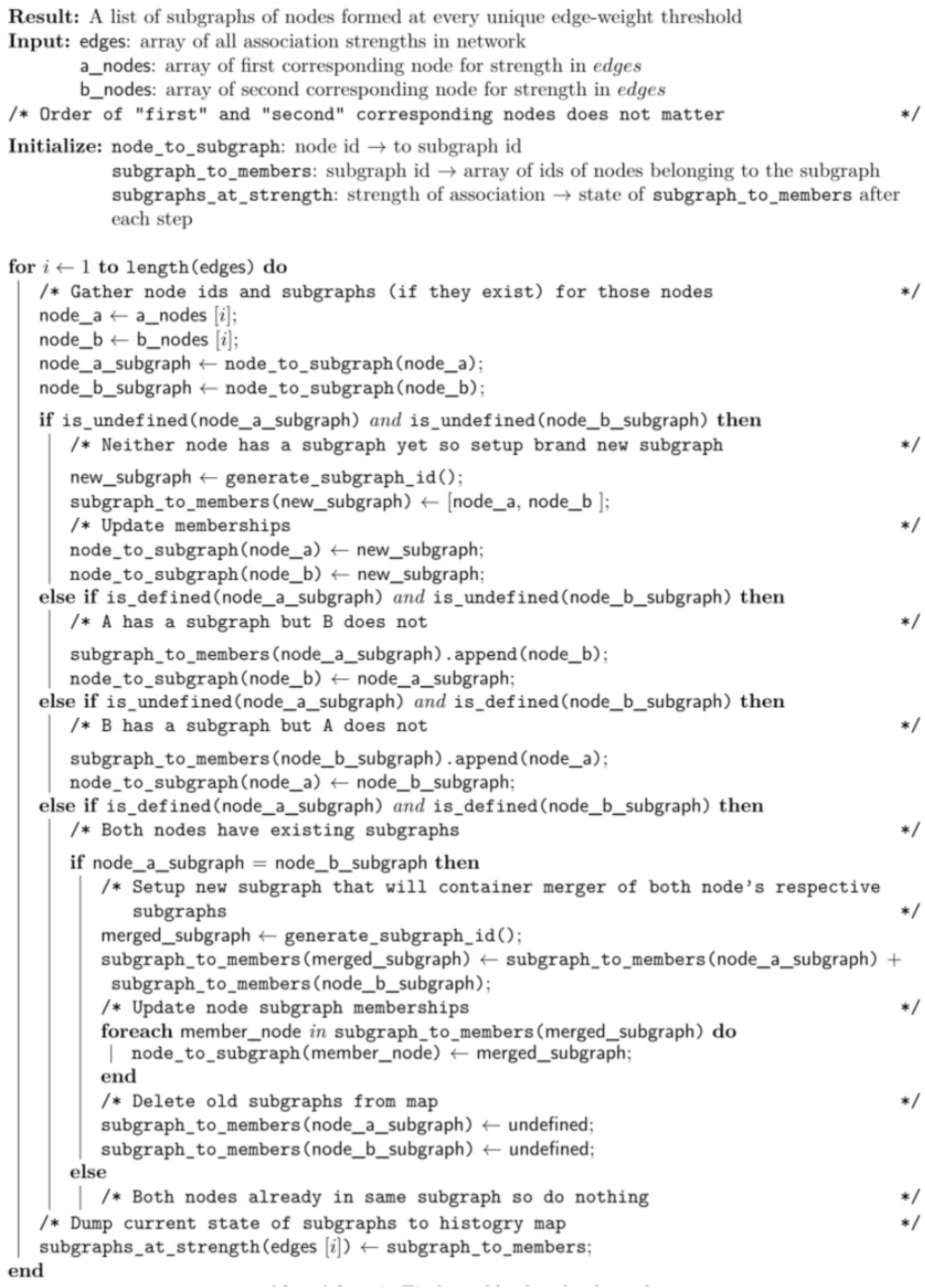

